# The measure and mismeasure of depression in older adults

**DOI:** 10.64898/2026.07.29.26359110

**Authors:** Malcolm Forbes, John J. McNeil, Michael Berk

**Affiliations:** The Institute for Mental and Physical Health and Clinical Translation (IMPACT), Deakin University, Geelong, Victoria 3220, Australia; School of Public Health and Preventive Medicine, Monash University, 553 St Kilda Road, Melbourne, Victoria 3004, Australia; Mental Health, Drugs and Alcohol Services, Barwon Health, University Hospital Geelong, Geelong, Victoria 3220, Australia

**Keywords:** Center for Epidemiologic Studies Depression Scale, CES-D-10, ASPREE, geriatric depression, late-life depression, screening, diagnosis, measurement

## Abstract

**Objectives:** The 10-item Centre for Epidemiological Studies Depression Scale (CES-D-10) has been used in cohort studies with scores of ≥ 8 and ≥ 10 used to indicate clinically significant symptoms or probable depression. We examined whether observations crossing these thresholds displayed core affective features expected in individuals who might be suffering with a clinically significant major depressive episode.

**Methods:** This was a descriptive analysis of repeated CES-D-10 assessments in the ASPirin in Reducing Events in the Elderly eXTension (ASPREE-XT) longitudinal cohort study of 19,114 participants. We identified observations of ‘probable depression’, defined as a CES-D-10 score ≥ 8 and then examined scores on item 3 (“I felt depressed”) and item 8 (“I was happy”). The primary outcome was the proportion of threshold-positive CES-D-10 observations with minimally depressed mood and preserved happiness.

**Results:** Over 13 years, 27,417 observations met the threshold of CES-D-10 ≥ 8. Of these, 13,356 observations (48.7%) had minimally depressed mood and preserved happiness.

**Conclusion:** A substantial proportion of observations considered ‘probable depression’ had little or no depressed mood, together with preserved happiness. CES-D-10 sum scores should be used with caution as a proxy for major depressive disorder in older adults.

## Introduction

There is an aphorism, variously attributed, that we only value what we can measure. For major depression, a serious medical condition affecting about 2% of older adults [1] and described as pain “quite unimaginable to those who have not suffered it” [2], measurement varies considerably. While a psychiatrist will usually spend an hour assessing a patient – considering their medical history, the context of their symptoms, and their degree of functional impairment [3] – this is not feasible in large research studies. Instead, depression rating scales are used, such as the Hamilton Rating Scale for Depression, Beck Depression Inventory, and Centre for Epidemiological Studies Depression Scale, as a proxy measure of the presence or absence of major depression. Concerns have been raised about the methodological and theoretical foundations of these rating scales, including inconsistent alignment between observer and self-reported rating scales, poor reliability, and rating scale development that prioritises ease of clinical use over validity [4].

Since its introduction over thirty years ago [5], the 10-item Centre for Epidemiologic Studies Depression Scale (CES-D-10) has been widely used in longitudinal cohort studies to measure depressive symptoms [6-8]. In the ASPirin in Reducing Events in the Elderly eXTension (ASPREE-XT) longitudinal cohort study, CES-D-10 was measured annually, with a total score of ≥ 8 out of 30 being used to classify probable depression [9].

For an individual to be diagnosed with major depression, they must experience either depressed mood or an inability to enjoy things for an extended period. Given concerns that the CES-D-10 may not correspond well to the definitions of major depression used in clinical practice, e.g. the International Classification of Diseases [10] and the Diagnostic and Statistical Manual [11], this study examined observations classified as representing probable depression based on their total CES-D-10 score, and estimated the proportion of these observations that did not have significant depressed mood and had preserved happiness, thus less likely to represent clinically significant or major depression.

## Methods

### Study design and sample

We conducted a secondary analysis of CES-D-10 scores for participants of ASPREE-XT, a cohort study of relatively healthy older American and Australian adults, described elsewhere [12]. We included those who had at least one CES-D-10 measurement. Most participants were followed-up over several years and thus contributed more than one CES-D-10 measurement.

### Measures

The CES-D-10 is a brief self-report measure of how often symptoms were experienced during the past week (Figure 1). Items are scored from 0 to 3 and summed to a total score from 0 to 30, with higher scores indicating more frequent symptoms. Item 3 is “I felt depressed” and is coded from 0 (“rarely or none of the time; less than 1 day”) to 3 (“all of the time; 5–7 days”). Item 8 asks “I was happy” and is reverse-coded, so that raw scores of 0 and 1 indicate feeling happy on 5–7 days and 3–4 days, respectively.

**Figure 1:**
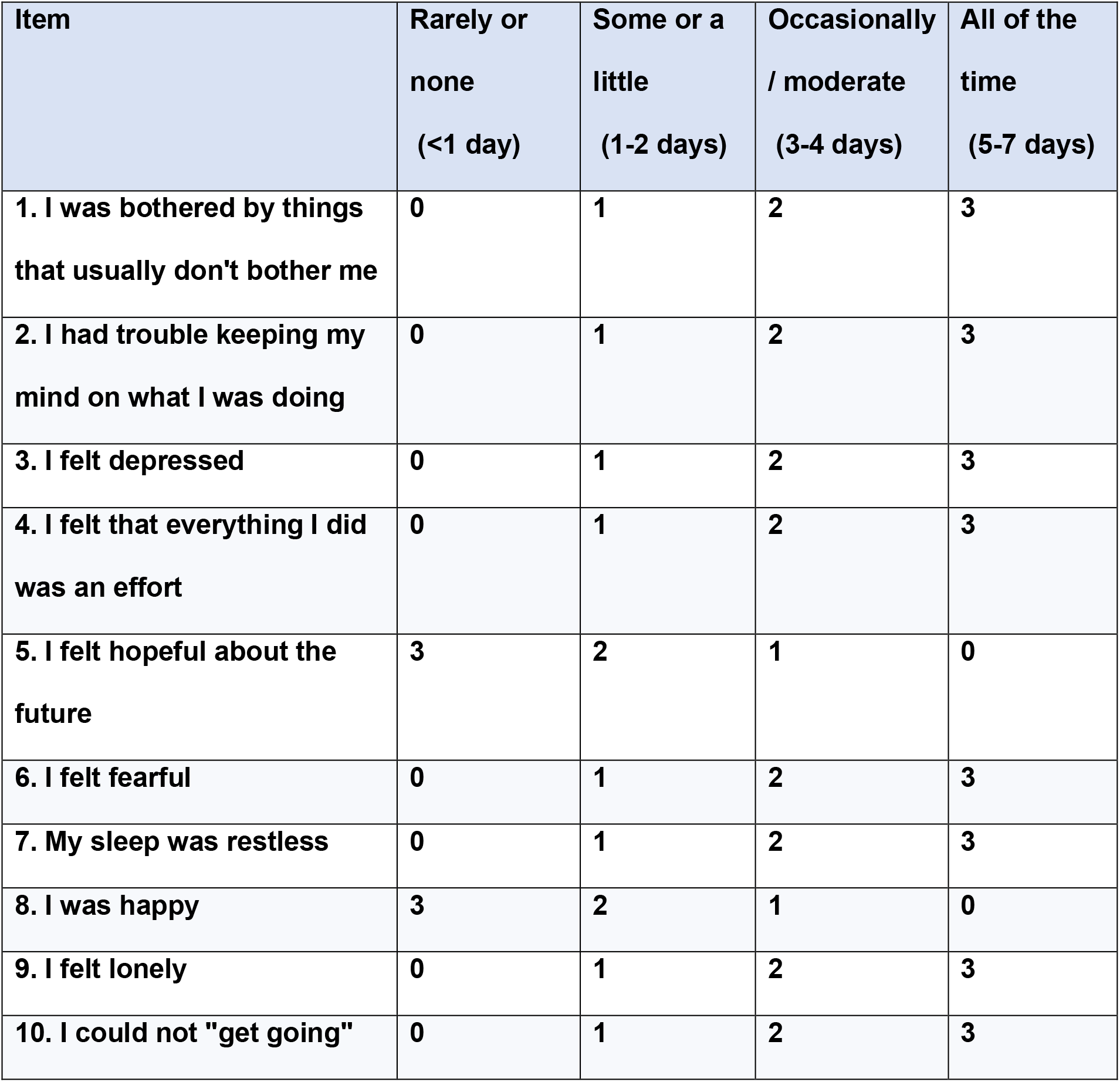
10-item Centre for Epidemiological Studies Depression Scale (CES-D-10)

We examined two CES-D-10 thresholds, ≥ 8 and ≥ 10, as both have been used widely in the late-life depression literature to suggest clinically significant depressive symptoms or probable depression [13,14]. We then examined two clinically relevant symptoms based on the requirements for a clinical diagnosis of depression – depressed mood and anhedonia. Minimal depressed mood was defined as a score of ≤ 1 on item 3 (see Figure 1), corresponding to feeling depressed on fewer than 3 days during the prior week. In a recent CES-D-10 psychometric analysis using data from our study, item 3 had the highest loading on the latent CES-D-10 factor [15], reinforcing its relevance as the scale’s clearest affective marker. Because the CES-D-10 has no direct anhedonia item, we used preserved happiness as an indicator of preserved positive affect rather than a direct measure of intact interest or pleasure. This was defined as a score of ≤ 1 on item 8 (see Figure 1), corresponding to feeling happy on 3–7 days during the prior week. The primary outcome was the proportion of observations of probable depression (defined as a CES-D-10 score of ≥ 8) where there was minimal depressed mood and preserved happiness. A secondary person-level analysis examined unique participants instead of observations.

We also selected a sample of individuals classified as depressed using their total CES-D-10 score and examined their linked demographic and clinical information within the ASPREE-XT dataset to explore contextual factors.

Finally, we examined hospital admissions with a primary diagnosis of depression as an indicator of severe clinically recognised depressive illness in the cohort.

## Results

### Observation-level results

With respect to individual observations of CES-D-10 defined probable depression (noting that an individual can have more than one depressive observation over the longitudinal study period), almost half of all CES-D-10 threshold-positive observations at ≥ 8 (13,356/27,417; 48.7%) occurred in participants reporting little or no depressed mood together with preserved happiness. At the stricter threshold of ≥ 10, more than one-third of observations (5,754/16,097; 35.7%) showed the same pattern. Table 1 shows the breakdown by each year, noting that CES-D-10 was not administered to all participants in year 2. Table 1 shows that the proportions remain relatively stable over time.

**Table 1:** Counts of threshold-positive CES-D-10 observations by year, and the subset with item 3 ≤ 1 and item 8 ≤ 1.

| **Year** | **Probable depression**<br>**(CES-D-10 $\geq 8$) (n)** | **Probable depression**<br>**(CES-D-10 $\geq 8$) without**<br>**depressed mood and**<br>**with preserved**<br>**happiness (n)** | **Percentage (%)** |
| --- | --- | --- | --- |
| 0 | 1879 | 978 | 52.0 |
| 1 | 2878 | 1412 | 49.1 |
| 2 | 1104 | 536 | 48.6 |
| 3 | 2938 | 1452 | 49.4 |
| 4 | 2581 | 1210 | 46.9 |
| 5 | 2679 | 1275 | 47.6 |
| 6 | 2532 | 1228 | 48.5 |
| 7 | 2363 | 1142 | 48.3 |
| 8 | 2310 | 1127 | 48.8 |
| 9 | 2224 | 1066 | 47.9 |
| 10 | 1847 | 903 | 48.9 |
| 11 | 1298 | 620 | 47.8 |
| 12 | 675 | 356 | 52.7 |
| 13 | 109 | 51 | 46.8 |
| **Overall** | **27417** | **13356** | **48.7** |

### Participant-level results

Overall, 6,953 of 9,581 participants (72.6%) who had a CES-D-10 score ≥ 8 at any point in the study had at least one threshold-positive observation with item 3 ≤ 1 (lack of significant depressed mood) and item 8 ≤ 1 (preserved happiness). The corresponding figure for CES-D-10 ≥ 10 was 3,811 of 6,818 (55.9%).

Five illustrative participants of the subgroup of interest are presented in Box 1, along with contextual information that may render a diagnosis of major depressive disorder less likely for each.

Over the course of follow-up, 9,581 participants (50.1% of all enrolled participants) had at least one CES-D-10 score ≥ 8. By contrast, hospital admission with a primary diagnosis of depression was uncommon, occurring in 121 participants (0.6% of all enrolled participants).

**Box 1: Individuals classified as having probable depression who may have an alternative explanation for their CES-D-10 score**

**Participant A** A woman in her mid-70s living alone at home reported no depressed mood but marked loneliness and restless sleep after her husband died; one year later, her CES-D-10 score had fallen from 9 to 3, suggesting possible grief and loneliness that resolved with the passage of time.

**Participant B** A man in his late 70s with a recently diagnosed haematological malignancy reported hopelessness and restless sleep, with a total CES-D-10 score of 11. His CES-D-10 score dropped to 4 one year later. His symptoms of hopelessness and poor sleep may be better explained by demoralisation associated with a cancer diagnosis rather than a major depressive disorder.

**Participant C** A man in his early 70s reported no depressed mood but marked effortfulness and poor concentration. Around the time of survey completion, his ferritin was very low, suggesting iron-deficiency as a contributor to his isolated fatigue and poor concentration, perhaps in lieu of a major depressive disorder.

**Participant D** A woman in her mid-70s reported no depressed mood, no loneliness, and preserved happiness, but endorsed effortfulness, restless sleep, and difficulty getting going most or all of the time. Her C-reactive protein level was significantly elevated around the time of survey completion, making a concurrent infection or inflammatory illness a plausible alternative explanation for her reported symptoms.

**Participant E** An man in his early 80s living at home with others who met the CES-D-10 threshold despite no depressed mood and preserved happiness. He had a seriously ill spouse and a significantly elevated thyroid-stimulating hormone concentration around the time of survey completion, suggesting plausible alternative biopsychosocial explanations for the elevated score.

## Discussion

For a diagnosis of depression, an individual must have either depressed mood or anhedonia contributing to significant functional impairment and there should not be a more plausible alternative explanation for their symptoms. Here we have shown that almost half of all observations of probable depression, defined as a CES-D-10 score of ≥ 8 out of 30, had preserved happiness without significant depressed mood. This makes a clinical diagnosis of depression less certain. As outlined in Box 1, contextual factors are important to consider as there is considerable overlap between non-specific symptoms of depression and symptoms present in a range of other medical conditions.

This study contributes to the body of literature that evaluates the use of depression rating scale sum-scores in depression measurement. Fried & Nesse [16] have synthesised research that suggests individual depressive symptoms are distinct phenomena which differ in their underlying aetiology, functional impact, and underlying biology.

Our findings have relevance for epidemiological and related research into the aetiology and management of depression, especially in the age of Big Data. If an outcome labelled probable depression is in fact an ill-defined distress phenotype, then exposure-outcome associations become difficult to interpret. A positive association of some putative risk marker may reflect predictors of loneliness, grief, poor sleep, physical illness burden, inflammation, pain, social adversity or demoralisation rather than predictors of major depression. A null association may be just as misleading, because any true disorder-specific signal may be diluted by the inclusion of many heterogeneous non-cases. As is often the case, this is best demonstrated with a clinical example. Few doctors would equate, (a) a woman experiencing sadness and insomnia following an adverse life event who recovers without intervention with, (b) a woman experiencing profound depressed mood, anhedonia, nihilistic delusions, and catatonia who requires and responds to involuntary electroconvulsive therapy. Methodological problems with depression caseness, as well as volunteer bias meaning that study participants are often non-representative of the general population, may be one reason why depression research has yielded few clinically important findings over several decades, despite billions of dollars of investment [17].

The study has limitations. First, we did not compare CES-D-10 responses with a structured psychiatric interview, so we cannot quantify diagnostic misclassification directly. Second, item 8 is not an anhedonia item, and preserved happiness is not identical to preserved interest or pleasure, thus individuals may have experienced some degree of anhedonia despite preserved happiness, although this would be rather unexpected. Finally, Gallo et al [18] suggest that depressed older adults may under-report explicit depressed mood. However, the presence of preserved happiness renders a putative non-dysphoric depression less convincing in this study. These limitations do not detract from the main point that a large body of late-life epidemiological research using CES-D-10-defined probable depression may in fact be studying an ill-defined distress phenotype rather than depressive disorder proper.

The physicist Paul Dirac remarked that the aim of science is “to make difficult things understandable in a simpler way” [19]. One of Dirac’s contemporaries clarified the point, noting that things should be made simple as possible but no simpler so as not to “surrender the adequate representation” [20]. In a laudable effort to better understand a debilitating disorder, academic psychiatrists and epidemiologists have tried to render something complex – major depressive disorder – more understandable in a simpler way by using brief, self-administered rating scales. Using the example of CES-D-10 in a longitudinal cohort study, we show that reducing major depressive disorder to a total score on a survey may not actually correspond to clinically meaningful or major depression diagnosed in the clinic.

Practising doctors should exercise discretion when reviewing the findings of research studies that have classified depressive symptoms or depression without considering contextual information.

## Data Availability

The aggregate results generated in this study are reported in the manuscript. Individual-participant data from ASPREE and ASPREE-XT are not publicly available because of participant privacy and consent restrictions. Data may be made available to qualified researchers upon reasonable request through the ASPREE Access Management System, subject to approval by the ASPREE Principal Investigators and completion of a data-sharing agreement.

